# Genome-wide gene by environment study of time spent in daylight and chronotype identifies emerging genetic architecture underlying light sensitivity

**DOI:** 10.1101/2022.06.29.22277078

**Authors:** Angus C. Burns, Andrew J. K. Phillips, Martin K. Rutter, Richa Saxena, Sean W. Cain, Jacqueline M. Lane

**Affiliations:** Turner Institute for Brain and Mental Health, School of Psychological Sciences, Monash University, Melbourne, VIC, Australia; Center for Genomic Medicine, Massachusetts General Hospital, Boston, MA, 02114, USA; Broad Institute, Cambridge, MA, USA; Division of Endocrinology, Diabetes & Gastroenterology, School of Medical Sciences, Faculty of Biology, Medicine and Health, University of Manchester, Manchester, UK; Diabetes, Endocrinology and Metabolism Centre, Manchester University NHS Foundation Trust, Manchester Academic Health Science Centre, Manchester, UK; Department of Anesthesia, Critical Care and Pain Medicine, Massachusetts General Hospital and Harvard Medical School, Boston, MA, 02114, USA; Division of Sleep and Circadian Disorders, Brigham and Women’s Hospital, Boston, MA, 02115, USA

**Keywords:** Light sensitivity, gene by environment, GWAS, chronotype, daylight, circadian rhythms, sleep, mood disorders, PTSD

## Abstract

Light is the primary stimulus for synchronizing the circadian clock in humans. There are very large interindividual differences in the sensitivity of the circadian clock to light. Little is currently known about the genetic basis for these interindividual differences. We performed a genome-wide gene-by-environment interaction study (GWIS) in 280,897 individuals from the UK Biobank cohort to identify genetic variants that moderate the effect of daytime light exposure on chronotype (individual time of day preference), acting as ‘light sensitivity’ variants for the impact of daylight on the circadian system. We identified a genome-wide significant SNP mapped to the *ARL14EP* gene (rs3847634; *p* < 5×10^−8^), where additional minor alleles were found to enhance the morningness effect of daytime light exposure (*β*_*GxE*_ = -.03, SE = 0.005) and were associated with increased gene *ARL14EP* expression in brain and retinal tissues. Gene-property analysis showed light sensitivity loci were enriched for genes in the G protein-coupled glutamate receptor signaling pathway and in *Per2*^+^ hypothalamic neurons. Linkage disequilibrium score regression identified significant genetic correlations of the light sensitivity GWIS with chronotype and sleep duration, such that greater light sensitivity was associated with later chronotype, greater insomnia symptoms and shorter sleep duration. Greater light sensitivity was also genetically correlated with greater risk for PTSD. This study is the first to assess light as an important exposure in the genomics of chronotype and is a critical first step in uncovering the genetic architecture of human circadian light sensitivity and its links to sleep and mental health.

## Introduction

In humans, daily rhythms in gene expression, physiology, and behavior are regulated by the circadian clock located in the suprachiasmatic nuclei^1^ (SCN). The circadian clock is primarily synchronized by ocular light exposure^2^. These rhythms are the overt manifestation of the activity of well-described molecular feedback-loops. Light information transmitted to the SCN from the retina via the retinohypothalamic tract (RHT) is translated into shifts in the phase of molecular biological clocks^3^ by altering the expression of core clock genes^4^.

There are large interindividual differences in the sensitivity of the human circadian clock to light. The circadian phase shifting response of humans to the same light stimulus can vary by as much as two hours^5^ and recent work has shown that the melatonin suppression response, a measure of circadian light sensitivity, can vary between individuals by over an order of magnitude^6^. These interindividual differences play a role in the aetiology of delayed sleep-wake phase disorder (DSWPD)^7^, major depressive disorder^8,9^, bipolar disorder ^10,11^ and other sleep disturbances^12^. Light sensitivity has also been related to tolerance of shift work, with individuals varying in their ability to adjust their biological clock so that their biological day aligns with the environmental night^13-15^.

Circadian light sensitivity is a heritable trait^16^, as measured by twin MZ-DZ correlations. Chronotype, a proxy-measure of circadian phase^17^, has significant SNP-heritability (SNP *h*^2^ range = .12-.37) in published GWAS and loci for this trait are enriched for circadian clock and glutamate signaling pathways^18,19^. These findings indicate that both circadian light sensitivity and the downstream traits it impacts are in part determined by genetic variation in the human population. A common variable-number tandem repeat polymorphism in the *PER3* gene has been associated with light sensitivity^20^ in a small sample; however, no other work has yet examined the genomics of circadian light sensitivity.

The timing of light exposure has a differential impact on the timing of circadian rhythms, as described by the human light phase-response curve (PRC). Light exposure in the late night and early day advances the rhythm while light exposure in the evening and early night delays the rhythm^21^. Consistent with the light PRC, daytime light exposure tends to produce advances in the phase of the circadian clock and thus is robustly associated with earlier chronotype and greater ease of awakening^22-24^.

Here, we leverage the phenotypic association between daytime light exposure and chronotype to conduct a Genome-Wide Interaction Study (GWIS) of loci that dampen or enhance this effect. Using unrelated individuals from the UK Biobank, we test for gene-by-environment (GxE) interactions between SNP alleles and daytime light exposure on chronotype to identify the genetic architecture of light sensitivity.

## Methods

### UK Biobank and genetic data

The UK Biobank prospective general population cohort contains more than 502,000 UK residents (aged 37–73 years; ∼54% women) recruited via National Health Service (NHS) patient registers from 2006 to 2010, with the study population described in detail elsewhere^25,26^. Participants provided extensive demographic, lifestyle, health, mood, and physical information via assessment and touch-screen questionnaires as well as physiological samples for the purpose of genome-wide array genotyping. Participants who accepted the invitation to join the UK Biobank cohort provided written, informed consent and the UK Biobank has generic ethical approval from the North West Multi-Center Research Ethics Committee (ref 11/NW/03820). The current analyses were conducted under UK Biobank application number 6818 (Martin Rutter).

In brief, blood, saliva, and urine were collected from participants, and DNA was extracted from the buffy coat samples. Participant DNA was genotyped on two arrays, Affymetrix UK BiLEVE Axiom array (initial 50,000 participants) and the Affymetrix UK Biobank Axiom Array with >95% common content and genotypes for ∼800,000 autosomal SNPs imputed to two reference panels (Haplotype Reference Consortium (HRC) and UK10K haplotype resource). Genotypes were called using Affymetrix Power Tools software. Sample and SNPs for quality control were selected from a set of 489,212 samples across 812,428 unique markers. Sample quality control (QC) was conducted using 605,876 high-quality autosomal markers. Samples were removed for high missingness or heterozygosity (968 samples) and sex chromosome abnormalities (652 samples). Genotypes for 488,377 samples passed sample QC (∼99.9% of total samples). Marker-based QC measures were tested in the European ancestry subset (n = 463,844), which was identified based on principal components of ancestry. SNPs were tested for batch effects (197 SNPs/ batch), plate effects (284 SNPs/batch), Hardy– Weinberg equilibrium (572 SNPs/ batch), sex effects (45 SNPs/batch), array effects (5417 SNPs), and discordance across control replicates (622 on UK BiLEVE Axiom array and 632 UK Biobank Axiom array; P value <10^−12^ or <95% for all tests).

For each batch (106 batches total), markers that failed at least one test were set to missing. Before imputation, 805,426 SNPs passed QC in at least one batch (>99% of the array content). Population structure was captured by principal component analysis on the samples using a subset of high-quality (missingness < 1.5%), high-frequency (>2.5%) SNPs (∼100,000 SNPs) and identified the subsample of white British descent. In addition to the calculated population structure by the UK Biobank, we locally further clustered participants into four ancestry clusters using K-means clustering on the principal components, identifying 453,964 participants of European ancestry. Related individuals were identified by estimating kinship coefficients for all pairs of samples, using only markers weakly informative of ancestral background. For the current analysis, individuals of non-white ethnicity and related individuals were excluded to limit confounding effects. The UK Biobank centrally imputed autosomal SNPs to UK10K haplotype, 1000 Genomes Phase 3, and Haplotype Reference Consortium (HRC). Autosomal SNPs were pre-phased using SHAPEIT3 and imputed using IMPUTE4. In total ∼96 million SNPs were imputed.. For all analyses, we used ∼15.0M HRC imputed variants with an imputation r2 ≥ 0.3, MAF ≥ 0.001 (0.1%) and with a Hardy–Weinberg equilibrium (HWE) P>1×10-12 (chi-squared; 1 degree of freedom). We excluded non-HRC imputed variants. Further details on the UK Biobank genotyping, quality control and imputation procedures can be found elsewhere^27^.

### Measurement of time spent in daylight, chronotype, and other self-reported phenotypes

At the baseline assessment, all participants self-reported how many hours they spent outdoors during the day on a typical day in both summer and winter (UK Biobank data-fields 1050 and 1060). Participants reported an integer using a touch-screen number pad or selected among alternate options including “Less than an hour a day”, “Do not know”, or “Prefer not to answer”. Initial cleaning of the data involved excluding participants that rated “Prefer not to answer” or “Do not know”, re-coding “Less than an hour a day” as zero (*n* = 98,431) and excluding values larger than the typical day length in the UK during summer (16 hours; *n* = 253) and winter (8 hours; *n* = 5,474). Summer and winter reports were strongly correlated (Pearson’s *r* = .65), indicating people who spent more time in outdoor light in winter also tended to do so in summer. Furthermore, a subset of approximately 20,000 participants completed a repeat assessment of this question from 2012-2013. Both summer and winter reports were strongly correlated (both *r* > .6) across repeat assessments, indicating good reliability of the measure. To yield a single measure of time spent in outdoor light, winter and summer reports were averaged within participants. For analysis, extreme bins (9-12 h spent outdoors) with low density (total *n* = 1,016 (0.2%) were collapsed into a single bin. Finally, due to the rightward skew in the distribution of the time spent in outdoor light, a log_10_ transformation was applied. We extracted self-reported chronotype from the baseline assessment (“Morningness-Eveningness”; data-field 1180) which was measured with the question “Do you consider yourself to be?” with six options: “Definitely a ‘morning’ person”, “More a ‘morning’ than ‘evening’ person”, “More an ‘evening’ than a ‘morning’ person”, “Definitely an ‘evening’ person”, “Do not know” or “Prefer not to answer”. The latter two categories were coded as missing. Participants further self-reported age and sex. Finally, weight and height were measured, and body-mass index (BMI) was calculated as weight (kg)/height^2^ (m^2^; UK Biobank data field 21001).

### Genome-wide association studies for light sensitivity in the UK Biobank

A genome-wide interaction study (GWIS) for circadian light sensitivity was conducted in unrelated individuals of white British ancestry (*n* = 280,897) to reduce confounding by stratification and kinship using the –gxe function in PLINK^28^. The logarithm of time spent in daylight (continuous) was used as the moderating exposure for the genotype effect on chronotype (continuous) with the following equation:

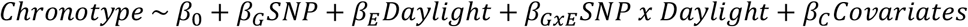

Here, SNP refers to the allelic dosage at a given SNP coded additively, daylight refers to the log-transformed average time spent outdoors during the day and the covariates included in the model were age, sex, genotyping array (UK BiLEVE or UK Biobank Axiom), BMI, and the top ten principal components of ancestry to control for population stratification. The interaction term (*β*_*GxE*_) was used to identify “light sensitivity” variants, or variants that moderate the strength of the effect of daylight exposure on chronotype. Negative values of *β*_*GxE*_ weights indicate additional minor alleles for a given SNP enhance the morningness effect of light on chronotype, whereas positive *β*_*GxE*_ weights indicate additional minor alleles dampen the morningness effect of light on chronotype.

In supplement to the GWIS, we conducted two separate GWAS of chronotype stratified by time spent in daylight to examine whether light exposure modifies the genetic architecture of chronotype. Low (≤1.5 h; *n* = 121,922) and high (≥3.5 h; *n* = 118,429) daytime light exposure groups were derived by applying a quartile split of the time spent in daylight variable, with the two intermediary quartiles excluded. Within these stratified samples, genetic association analysis across the autosomes was performed in European participants with BOLT-LMM^29^ linear mixed models allowing for relatedness using an additive genetic model adjusting for age, sex, ten principal components of ancestry, genotyping array, and genetic correlation matrix.

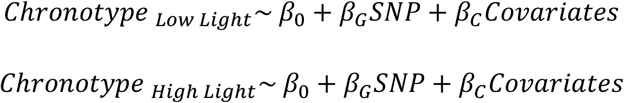

A maximum SNP missingness of 10% and per-sample missingness of 40% was tolerated. A SNP imputation quality threshold of 0.80 and minor allele frequency of 0.001 (0.1%) was used for all genome-wide analyses. A genome-wide significance threshold of 5×10^−8^ was used to identify significant SNPs.

### Gene, Pathway, Tissue, and Single-Cell Expression

Post-GWIS gene based and pathway analyses were carried out with MAGMA^30^ in FUMA^31^. Gene based analysis was performed using default MAGMA parameters and a SNP-wide mean model of SNPs mapped to 18,960 protein coding genes (Bonferroni *p* = 0.05/18960 = 2.64×10^−6^). Pathway analysis was conducted using MAGMA^30^ gene-set analysis in FUMA, which tests 10,894 gene sets (curated gene sets = 4,728; gene ontology terms = 6,166) for enrichment using the full distribution of SNP *p* values from the GWIS (Bonferroni *p* = 0.05/10,651 = 4.69×10^−6^). Tissue enrichment analyses (GTEx V8) were conducted using MAGMA gene-property analysis in FUMA using default parameters to test the relationship between tissue-specific gene-expression profiles and the GWIS light sensitivity associations. Cell-type specificity analysis was conducted in the same manner, using single-cell RNA-seq gene-expression profiles as the outcome, on the three circadian hypothalamic cell subtypes (*Per2*^*+*^, *VIP*^*+*^ & *NMS*^*+*^) identified in Romanov, Zeisel, Bakker, Girach, Hellysaz, Tomer, Alpár, Mulder, Clotman, Keimpema, Hsueh, Crow, Martens, Schwindling, Calvigioni, Bains, Máté, Szabó, Yanagawa, Zhang, Rendeiro, Farlik, Uhlén, Wulff, Bock, Broberger, Deisseroth, Hökfelt, Linnarsson, Horvath and Harkany ^32^. Queries of expression, methylation and histone modification QTLs for genome-wide significant loci were done using QTLbase^33^ and EyeGEx^34^.

### Genetic Correlations

Linkage Disequilibrium (LD) score regression was implemented in the R package *GenomicSEM*^*35*^ to examine the genetic correlation (*r*_g_) between GWIS light sensitivity genetic architecture (indexed by *β*_*GxE*_) and sleep, circadian, psychiatric, and metabolic traits with publicly available GWAS data that have been linked phenotypically to circadian light sensitivity^36-39^. A second set of genetic correlations was conducted for stratified GWAS of chronotype in high and low daylight exposure groups and the above traits to examine whether the exposure moderated the association of chronotype with these traits. Genetic correlations for high and low daylight exposure chronotype GWAS were compared using a χ^2^ test of a constrained 1-df model, where the genetic correlation of chronotype in the high and low daylight groups was constrained to be equal against an unconstrained 0-df model in *GenomicSEM*.

## Results

### Phenotypic results

The median time spent in daylight among UK Biobank participants was 2.5 h (IQR = 1.5-3.5 hours) and time spent in daylight was correlated with chronotype, such that greater daylight exposure was associated with greater morningness (*r* = -.10, *p* < 5×10^−150^). This association has been previously reported^24^ and is independent of lifestyle, demographic, and employment covariates. Demographic characteristics of the GWIS sample are presented in Supplemental Table 1.

### Genome-wide interaction study

The Manhattan and Q-Q plots for the GWIS of light sensitivity SNPs using 14,055,103 imputed variants in unrelated white British UK Biobank participants (*n* = 280,897; λGC = 1.04) are presented in Figure 1a-b. A single genome-wide significant hit, rs3847634 (see Supplemental Figure 1 for locus plot), was identified that mapped to the *ARL14EP/FSHB* genomic region on chromosome 11 (*chr11:30,344,598-30,359,774*), *β*_*GxE*_ = -.03, SE = 0.005, *Z* = -5.48, *p* = 4.37 ×10^−8^), such that additional T allelic dosage conferred a stronger morningness effect of daylight (see Figure 1c; see Supplemental Table 2 for top GWIS hits). Sensitivity analysis that removed the adjustment for BMI replicated this finding (*β*_*GxE*_ = -.03, *p* = 4.5×10^−8^). The marginal effect of this SNP in the GWIS analysis was positive and genome-wide significant (*β*_*G*_ = .04, SE = .007, *Z* = 6.42, *p* = 1.35 ×10^−10^), indicating that, independent of daytime light exposure, additional T alleles conferred greater eveningness. Importantly, this marginal effect was not identified in a prior GWAS of chronotype^18^ (rs3847634 β = 0.01, SE = 0.003, *p* = 0.0004) that did not take into account daytime light exposure and its interaction with genotype. An independent *ARL14EP/MPPED2* locus (rs621421; *r*^2^ with rs3847634 = .008, 1000 Genomes EUR population) has been associated with chronotype^18^ in prior GWAS; however, neither the marginal nor interaction effect for this locus reached significance in the GWIS (*p*_both_ > .05). The *p* values for the marginal effect of SNPs on chronotype in the GWIS and the published chronotype GWAS^18^ effect correlated strongly (*r* = .56) and the genetic correlation between the two indicated agreement (*r*_g_ = 1.01). The GxE effect from the GWIS for all previously reported chronotype loci are reported in Supplemental Table 3. No other novel genome-wide significant marginal effects were identified in the GWIS as compared to the prior chronotype GWAS.

**Figure 1.**
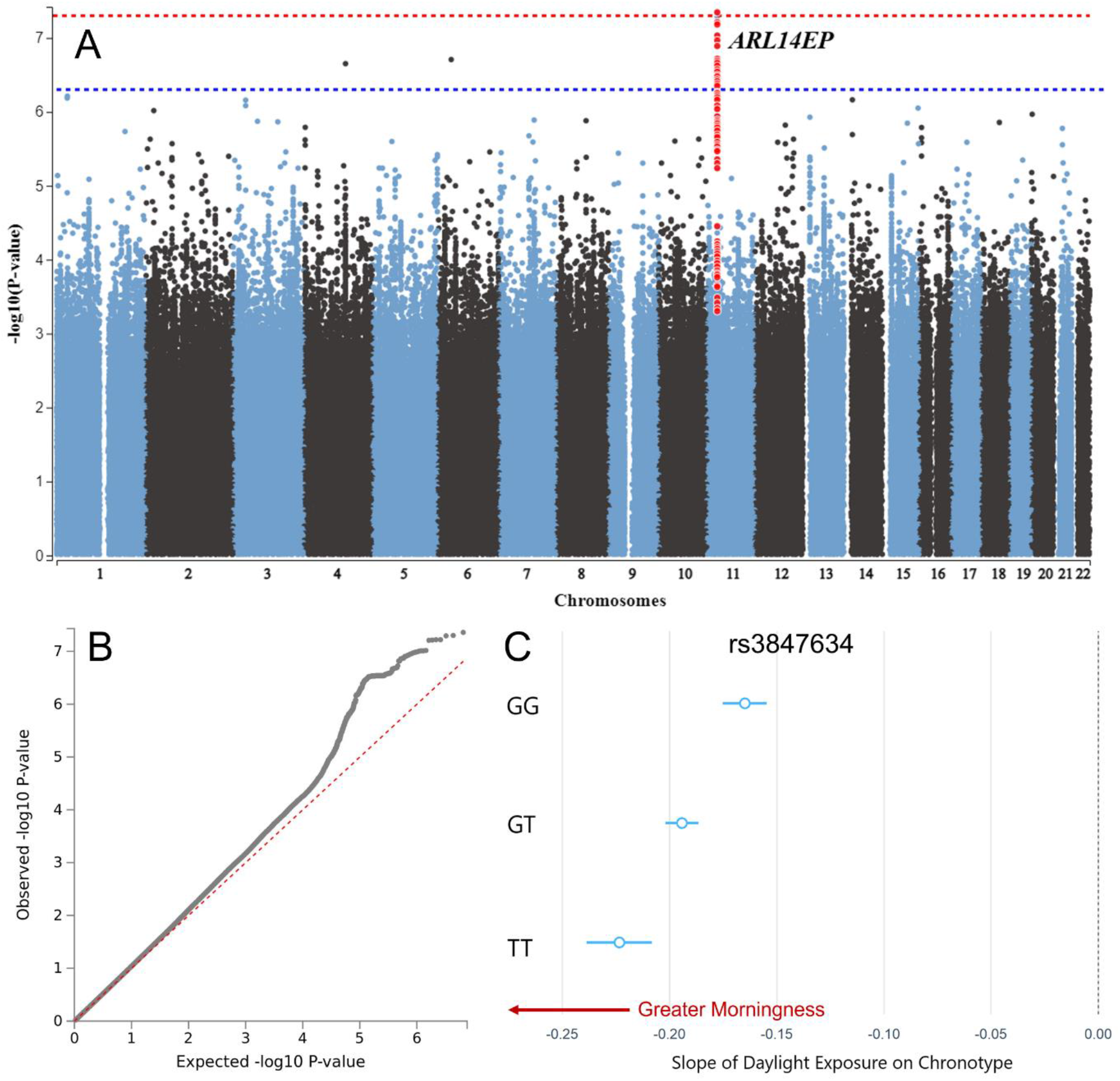
Genome-wide interaction study of light sensitivity. (A) Manhattan and (B) Q-Q plots of genome-wide interaction study (GWIS) for daylight exposure on chronotype, horizontal red dotted line indicates genome-wide significance threshold p < 5×10^−8^. The genome-wide significant SNP rs3847634 at the *ARL14EP* locus and SNPs in LD with it are highlighted in red (C) Conditional regression plot demonstrating the interaction between genotype at rs3847634 and daylight on chronotype, such that additional T alleles confer a stronger morningness effect of daylight.

### QTL, Gene, and Pathway Analysis

The rs3847634 SNP is an expression quantitative trait locus (eQTL; QTLbase & EyeGEx) for *ARL14EP* in the hippocampus (β = .061, *p* = 1.15×10^−7^) and retina (β = .44, *p* = 5.49×10^−10^), such that additional T alleles were associated with increased *ARL14EP* expression in these tissues. It was also associated with increased *FSHB* gene expression in the brain (β = .25, *p* = 5.23×10^−12^; Supplemental Tables 4 & 5). Analysis of histone and methylation QTLs revealed that the T allele is associated with increased H3K27ac histone modifications (β = .06, *p* = 2.52×10^−6^) and reduced methylation (cg06241208; chr11:30,344,200; β = -.34, *p* = 3.76×10^−16^; Supplemental Table 6) at the promoter region of the *ARL14EP* gene.

MAGMA gene-based tests combining SNP-level information within a gene did not identify any Bonferroni-significant genes (see Supplemental Figure 2; Supplemental Table 7). However, suggestive signals were identified in the *HOMER1* (*p =* 1.37×10^−5^), *GOLIM4* (*p* = 3.34×10^−5^), and *ARL14EP* (*p* = 4.43×10^−5^) genes. Pathway analysis combining gene-level information identified a single Bonferroni-significant Gene-Ontology pathway, the G Protein-Coupled Glutamate Receptor Signaling Pathway (*p* = 1.87×10^−6^; see Supplemental Table 8), which contains 13 genes, including *HOMER1* and the glutamate metabotropic receptor 1 gene (*MgluR1/GRM1*; Gene-based test *p* = 0.006; see Supplemental Table 8). Additionally, genes identified in the light sensitivity GWIS by QTL and positional mapping (RGS12 & RIC8B) significantly overlapped with the Gene Ontology *G Protein Alpha Subunit Binding* (*p*_adj_ = .01; see Supplemental Figure 3).

### Tissue and Single-Cell Expression Analysis

MAGMA tissue-expression analysis did not identify significant enrichment in any of the 53 superordinate GTEx tissue types (Supplemental Figure 4), nor any of the brain samples across the stages of development in the BrainSpan database (Supplemental Figure 5). Single-cell RNA-seq gene-property analysis of circadian subtypes of hypothalamic neurons^32^ revealed a Bonferroni-significant enrichment in *Per2*^+^ hypothalamic cells (*p* = 0.007; Bonferroni *p* = 0.017), but not in *VIP*^+^ or *NMS*^+^ circadian subtypes (Supplemental Table 9).

### Genetic Correlations

We performed LD-score regression^40^ (LDSC) analyses to assess the genetic overlap of light sensitivity (i.e., the genetic architecture of loci moderating the effect of daylight on chronotype) and publicly available sleep, circadian, psychiatric, and metabolic traits (see Figure 2 and Supplemental Table 10). Light sensitivity (*h*^2^ = .2%, *se* = .1%) had a strong negative genetic correlation with chronotype (*r*_g_ = - .62, se = 0.10, *p* = 1.47×10^−9^), indicating that SNPs which tend to confer greater light sensitivity (i.e., enhancing the morningness effect of daylight on chronotype) tend to also be eveningness SNPs. Sleep duration also significantly correlated with light sensitivity (*r*_g_ = .35, se = 0.11, *p* = 0.002), indicating that SNPs that tend to confer lower light sensitivity tend to also confer longer sleep duration. There was a negative genetic correlation between light sensitivity and PTSD (*r*_g_ = -.71, se = 0.30, *p* = 0.01), indicating that SNPs associated with higher light sensitivity tended to confer greater risk for PTSD; however, the uncertainty for this estimate was large. Similarly, frequent insomnia had a significant genetic correlation with light sensitivity (*r*_g_ = -.26, se = 0.12, *p* = .03), such that greater light sensitivity was associated with greater insomnia symptomology. No other traits were significantly genetically correlated with light sensitivity.

**Figure 2.**
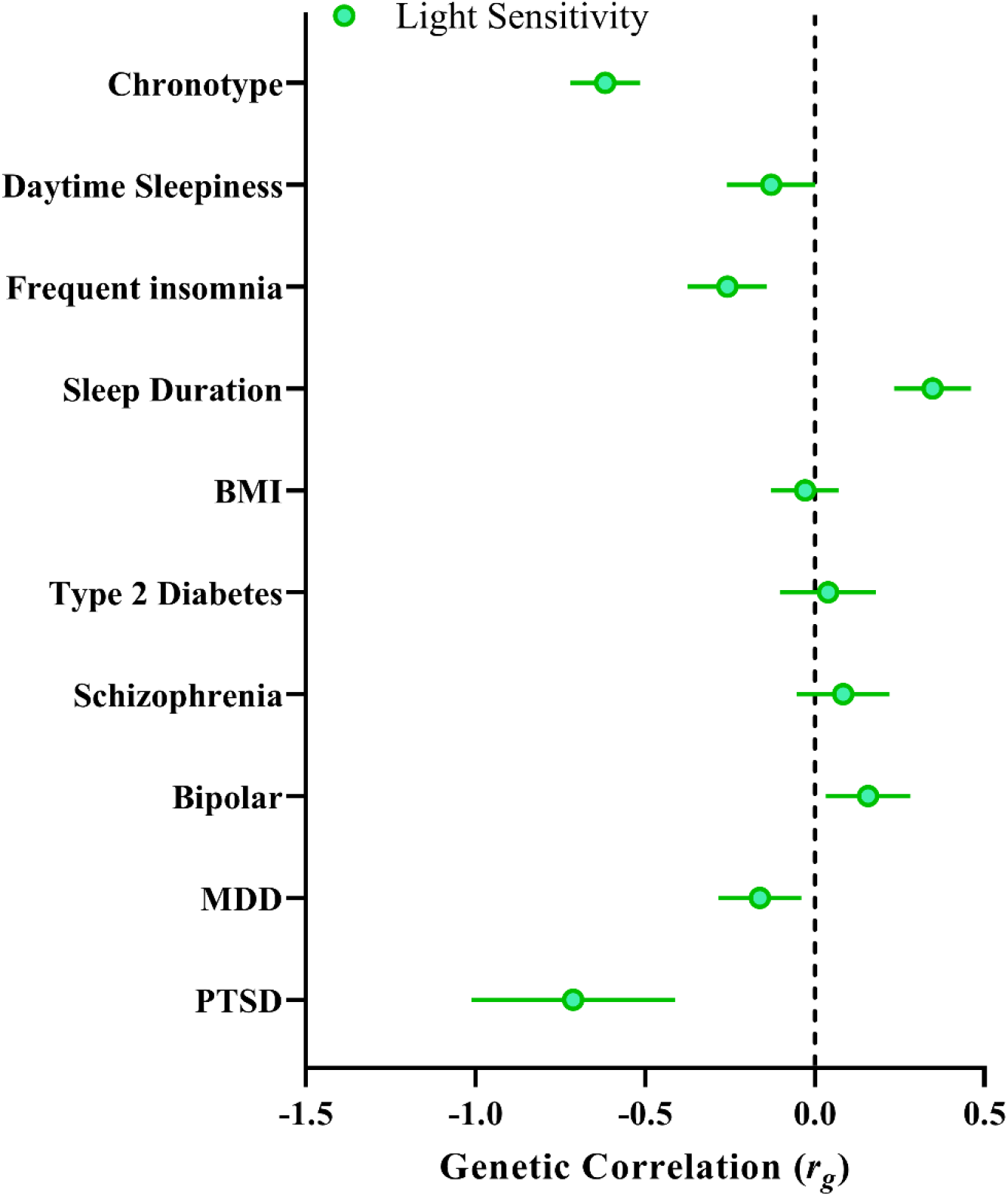
Light sensitivity genetic correlations. Genetic correlations between the light sensitivity GWIS and sleep, psychiatric and metabolic traits previously linked to light sensitivity. Note: as negative values of the β_GxE_ term indicate an enhancing effect of the SNP on light, negative genetic correlations denote the correlation between greater light sensitivity with a given trait.

We then tested if the amount of daytime light exposure moderates the genetic correlations of chronotype with the same set of sleep, circadian, psychiatric, and metabolic traits by performing LDSC on chronotype GWAS stratified by daylight exposure (bottom quartile, low light ≤ 1.5h, *n* = 121,922; top quartile, high light ≥ 3.5h, *n* = 118,429; see Supplemental Figures 6 & 7 for Manhattan and Q-Q plots). The SNP-*h*^2^ of chronotype was moderated by light exposure as calculated by BOLT-REML, with heritability being higher in the low daylight exposure group (Chronotype_Low Light_ *h*^2^ = 0.20, se = 0.003; Chronotype_High Light_ *h*^2^ = 0.16, se = 0.003). The genetic correlation between chronotype in those exposed to low and high daytime light was strong (*r*_g_ = 0.93, se = 0.05, *p* = 6.92×10^−88^). The lead SNP of the low light chronotype GWAS was rs12424741, mapped to the ALG10 locus (*p* = 5.29×10^−16^) and the lead SNP of the high light chronotype was rs3769124 mapped to the ASB1 locus (*p* = 2.6×10^−10^), both previously reported^18^. Consistent with the light sensitivity genetic correlations, there was a significant difference between the genetic correlation of chronotype and sleep duration in the high (*r*_g_ = 0.11, se = 0.03, *p* = 1.5×10^−4^) versus low (*r*_g_ = 0.02, se = 0.03, *p* = 0.49; χ^2^ = 6.06, *p* = 0.01) light exposure groups, such that eveningness was associated with longer sleep duration in those exposed to high daytime light but not in those exposed to low daytime light (see Figure 3 and Supplemental Table 11). Similarly, the genetic correlations between chronotype and PTSD (χ^2^ = 4.81, *p* = 0.03) and chronotype and daytime sleepiness (χ^2^ = 4.12, *p* = 0.04) were significantly different, such that eveningness was correlated with reduced risk for PTSD and daytime sleepiness among those exposed to high levels of daytime light. Eveningness was more strongly associated with major depressive disorder among those with low daylight exposure (*r*_g_ = 0.10, se = 0.03, *p* = .0004) compared to those with high daylight exposure (*r*_g_ = 0.06, se = 0.03, *p* = .03), though this difference was only suggestive (χ^2^ = 2.99; *p* = .08).

**Figure 3.**
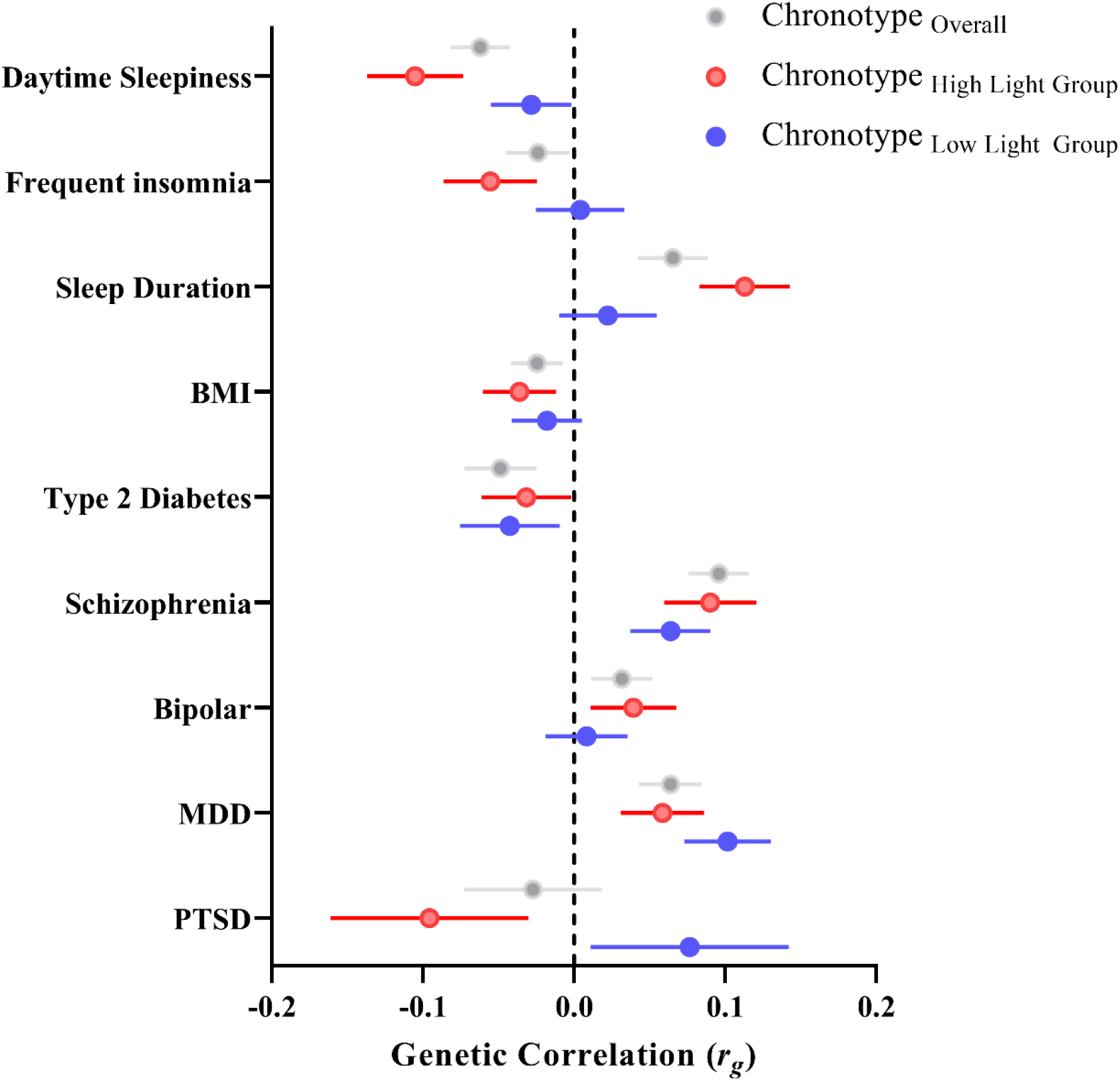
Chronotype genetic correlations stratified by daylight exposure. Genetic correlations between chronotype (eveningness) and sleep, psychiatric and metabolic outcomes for GWAS split by environmental exposure to daylight (bottom quartile, blue points, low light ≤ 1.5h, *n* = 121,922; top quartile, red points, high light ≥ 3.5h, *n* = 118,429). Grey points depict the genetic correlations of the published chronotype GWAS with sleep, psychiatric and metabolic outcomes for comparison.

## Discussion

Here we report the first study of genome-wide gene by environment interactions for daytime light exposure on chronotype and uncover an emerging genetic architecture of human circadian light sensitivity. We identified a novel locus, candidate genes, and pathways for the modulation of the advancing effect of daytime light exposure on chronotype. We also observed enrichment for light sensitivity genes in hypothalamic *Per2*^+^ cell subtypes. Genetic correlations indicated that the genetic architecture of light sensitivity significantly overlapped with chronotype and sleep duration, while being nominally associated with sleep and psychiatric disturbance. This study provides important insights into the biological basis of human light sensitivity and its link to sleep and psychiatric traits.

We found that one locus, rs3847634, mapped to the *ARL14EP* gene on chromosome 11, reached genome-wide significance in the GWIS. Additional T alleles at this locus were associated with a stronger morningness effect of daytime light exposure on chronotype, thus conferring enhanced light sensitivity. In addition to identifying this GxE effect, we observed that the rs3847634 SNP reached genome-wide significance for its marginal effect on chronotype, such that additional T alleles at this locus were associated with greater eveningness, an effect previously unobserved^18,19^. An independent SNP in the *ARL14EP* gene (rs621421, *r*^2^= 0.0075 with rs3847634) that has previously been associated with chronotype^18^, provides additional evidence for this gene playing a role in circadian timing. This SNP did not interact with light in the present study. Replication of this interaction in independent datasets with light exposure, chronotype and genetic data will be important.

This result highlights the importance of considering relevant environmental exposures when examining the genomics of a given trait. While potentially counterintuitive, this result is consistent with a variant that enhances the overall effect of light on the timing of the circadian clock, independent of the timing of light exposure. Enhanced sensitivity to light would boost phase-advancing effects of daytime light exposure, thereby strengthening the association between daylight exposure and morning chronotype. However, the same enhancement of light sensitivity would also boost phase-delaying effects of evening light exposure^41^. Given modern human lighting conditions are characterized by reduced bright light exposure during the day and bright electric light exposure at night^42-45^ a SNP that enhances light sensitivity would therefore be expected to confer both of these properties.

We found that rs3847634 is associated with reduced CpG methylation and increased h3k27ac histone modifications in the promoter region of the *ARL14EP* gene. Such epigenetic modifications are consistent with a more active gene promoter by diminishing the repressive effects of DNA methylation and opening chromatin to transcriptional machinery, respectively^46,47^. These modifications may therefore explain our observation that rs3847634 is an eQTL for increased *ARL14EP* gene expression in brain and retinal tissues. Together, these findings hint at a plausible molecular mechanism for the effect of rs3847634 on *ARL14EP* gene expression and subsequent phenotypic effects.

The *ARL14EP* gene is important for appropriate brain and axonal development. In humans, *ARL14EP* haploinsufficiency is associated with callosal hypoplasia while its knockdown in animal models results in severe axonal arborization deficits and the disruption of transcollosal connectivity^48^. WAGR syndrome (Wilms’ tumour, Aniridia, Genital abnormalities, and mental Retardation) is caused by an 11p14-12 deletion that includes the *ARL14EP* gene and is characterized by a photosensitivity and intellectual disability phenotype^49^. Furthermore, an independent SNP at the *ARL14EP* locus is associated with retinal refractive error^50^ and non-synonymous mutations in the CRD domain of the ARL14EP protein are associated with neurodevelopmental disorders^51,52^. There appears to be overlap in biological function of *ARL14EP* with other genes that encode for axon development/guidance (via semaphorins, ephrins). The semaphorin Sema6a is hyper-expressed in *ARL14EP*-knockdown neurons and epigenetic editing of Sema6a gene promoters has been shown to rescue axon function in *ARL14EP* knockdown animal models^48^. Semaphorins and ephrins have been shown to be important in the development of retinal ganglion cells and a SNP at the Sema6a locus is associated with chronotype^18,53,54^. While indirect, collectively these data in concert with our findings point toward a putative role for *ARL14EP* in axonal development pertinent to the retina and phototransduction – for example, by altering retinohypothalamic tract development and subsequently the strength of retinal inputs to the circadian clock.

Pathway analysis identified enrichment of GWIS SNPs in the G protein-coupled glutamatergic signalling pathway. Nominal enrichment at the gene level was also observed for the *HOMER1* and *MgluR1* genes that are constituents of this pathway. These results are consistent with a large body of research that demonstrates the critical role of glutamatergic signalling in circadian photoreception, in particular via metabotropic glutamate receptors 1 and 5 expressed in the SCN^55,56^. Similarly, there is strong evidence for the role of HOMER1 in circadian phototransduction. HOMER1 participates in activity-dependent control of metabotropic glutamate receptors, including MgluR1 and MgluR5, is rhythmic in the SCN with a peak in the subjective night when the clock is most sensitive to phase shifts, and its expression is induced by light exposure in the subjective night^57-59^. Furthermore, cell-type specificity analysis focusing on hypothalamic neurons expressing circadian-related genes^32^ identified significant enrichment in SCN neurons expressing the core clock gene Per2. Together, these findings provide convergent evidence towards a circadian light sensitivity understanding of the genetic architecture uncovered by the present GWIS.

Finally, genetic correlation analyses identified significant genetic overlap between light sensitivity and chronotype, such that greater light sensitivity was associated with greater eveningness. This genetic correlation was strong and is likely explained by a similar phenomenon described above, whereby modern human lighting environments are depleted for daytime light^43^ and enriched with night-time light^44,60^ – hence resulting in light sensitivity SNPs being associated with eveningness as individuals with a genetic load for greater light sensitivity experience greater delaying phase shifts to evening light and a later chronotype^21,61^. Importantly, there is not a statistical dependence between the light sensitivity GxE term in the model and the marginal SNP effect. For example, a recent stress-sensitivity GWIS for depression found zero genetic correlation between stress sensitivity and the marginal SNP effect on depression^62^.

Light sensitivity was also found to genetically correlate with sleep duration, such that greater light sensitivity genetic load was associated with lower sleep duration genetic load. This finding is consistent with greater light sensitivity amplifying the phase delaying effect of evening light exposure, resulting in the delayed timing of sleep and, given a constant wake time, shortened sleep^12,44^. Greater light sensitivity may also shorten sleep via direct effects on arousal^63,64^, thereby enhancing the disruptive effects of evening light on subsequent sleep^65^. A novel association between greater light sensitivity and PTSD diagnosis was also observed. This finding may elucidate recent work demonstrating circadian dysrhythmia is linked to symptom severity in PTSD and that melatonin and phase advancing light may be effective treatments for PTSD symptoms^66-68^.

We note that there are limitations of both the sample and the subjective nature of predictors and outcome measures used in this study. Firstly, there are significant changes in the properties of the circadian system that occur with age^69^, including earlier phase^70^, dampened amplitude^70^ and reduced light sensitivity^71-73^. Older adults also spend a larger portion of their day in bright (>1000lux) light^74^. Given these changes and the makeup of the UK Biobank sample being largely middle-older aged adults, it is feasible that the genetic architecture of light sensitivity, defined here as the moderating effect of genotype on the daylight-chronotype relationship, may differ in younger adults. Secondly, the present work relies on a self-report measure of daytime light exposure. While this measure may broadly capture time spent in bright light it does not capture objective intensity and duration of exposure at the ocular level. Future work should address these limitations by testing examining genetic light sensitivity across the lifespan using objective measures of light and, if feasible, circadian phase. Finally, it is important to note that selective gene-environment correlation may provide an alternate explanation for our results, such that individuals with a greater genetic propensity to seek out more daylight may also therefore tend to have earlier chronotype.

The present work is the first to identify common genetic variation underlying light sensitivity in the human genome; its links to molecular pathways and cell subtypes in the hypothalamus; and genetic overlap with sleep duration, sleep disturbance and PTSD. This study emphasises the relevance of light as a critical environmental exposure when studying the genomics of circadian rhythms traits and provides the basis for future lines of research to improve the methodology beyond subjective measures of light and circadian phase.

## Data Availability

All data produced in the present study are available upon reasonable request to the authors

## Acknowledgements

This research and Angus C. Burns was supported by a Research Training Program (RTP) scholarship from the Australian Government.

## Author Contributions

*Study conceptualisation and design:* A.C.B., J.M.L., R.S., S.W.C., A.J.K.P., M.K.R.; *Analysis:* A.C.B., J.M.L.; *Overseeing project:* A.C.B., J.M.L., R.S., S.W.C., A.J.K.P.; *Writing and editing:* A.C.B., J.M.L., R.S., S.W.C., A.J.K.P., M.K.R.; *Critical revisions of the manuscript:* A.C.B., J.M.L., R.S., S.W.C., A.J.K.P., M.K.R.

## Data Availability Statement

This work utilized the UK Biobank resource (application 26209; Martin Rutter).

