## Supplementary Figures for "Genome-wide gene by environment study of time spent in daylight and chronotype identifies emerging genetic architecture underlying light sensitivity"

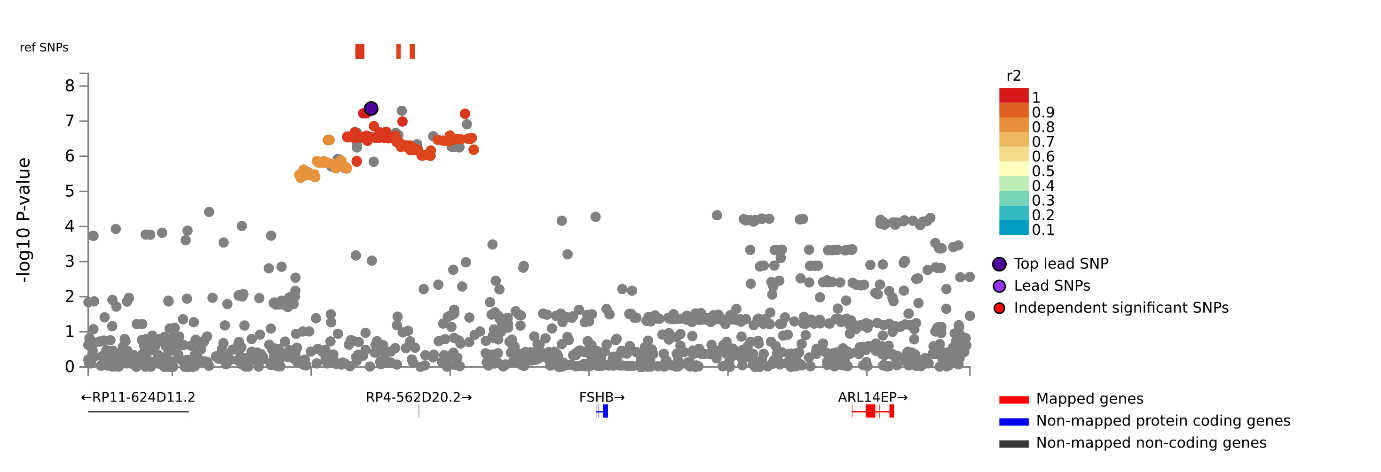

**Supplementary Figure 1.** Locus-zoom plot from FUMA depicting the location of the rs3847634 SNP mapped positionally and functionally via eQTLs to the *ARL14EP* gene.

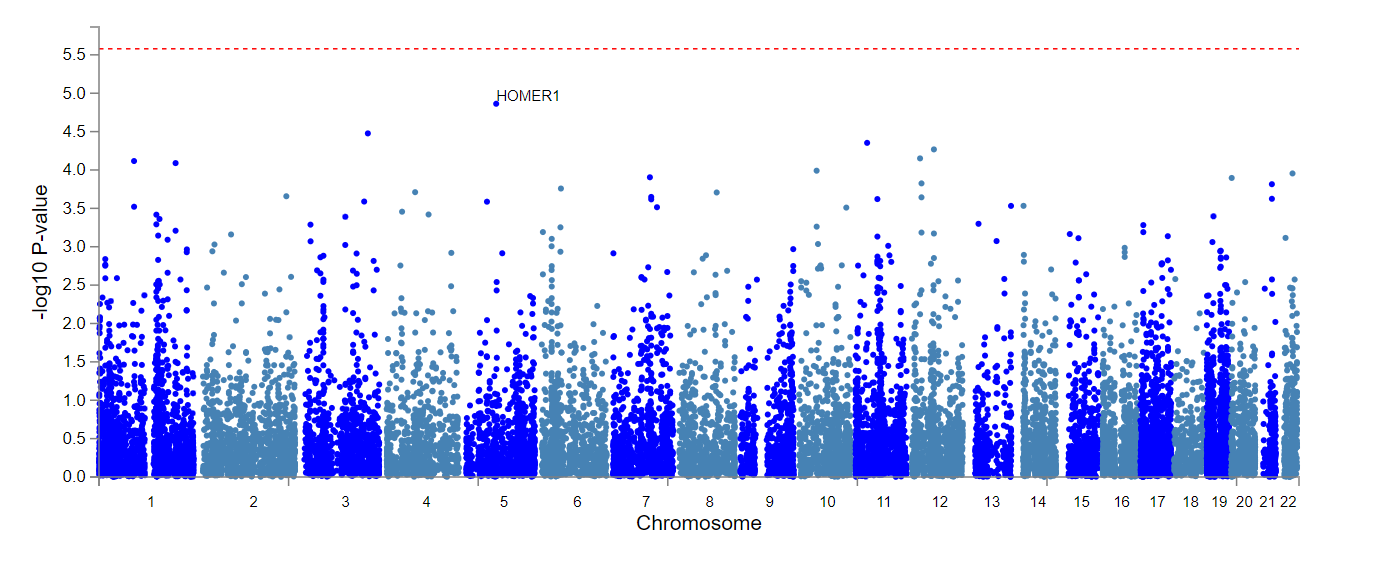

**Supplementary Figure 2.** Manhattan plot of MAGMA gene-based tests of GWIS.

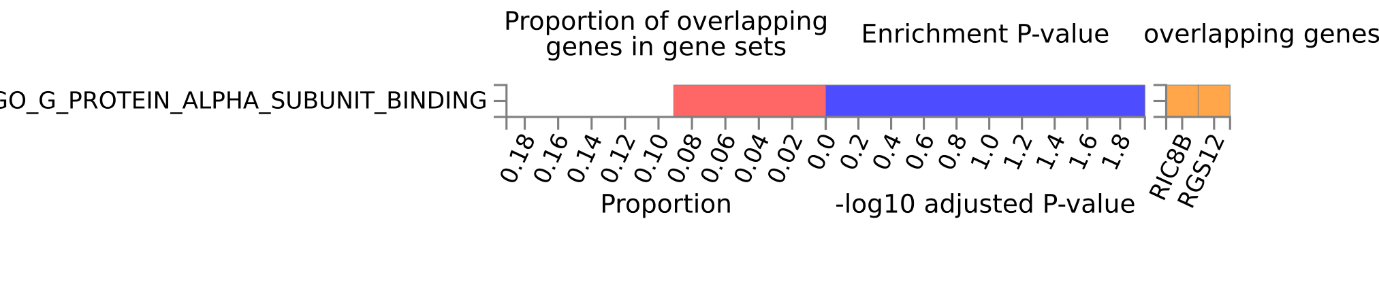

**Supplementary Figure 3.** Significant enrichment of G protein alpha subunit binding pathway identified by FUMA GENE2FUNC differentially expressed genes (DEG) analysis of prioritised genes identified by positional mapping analysis of GWIS loci.

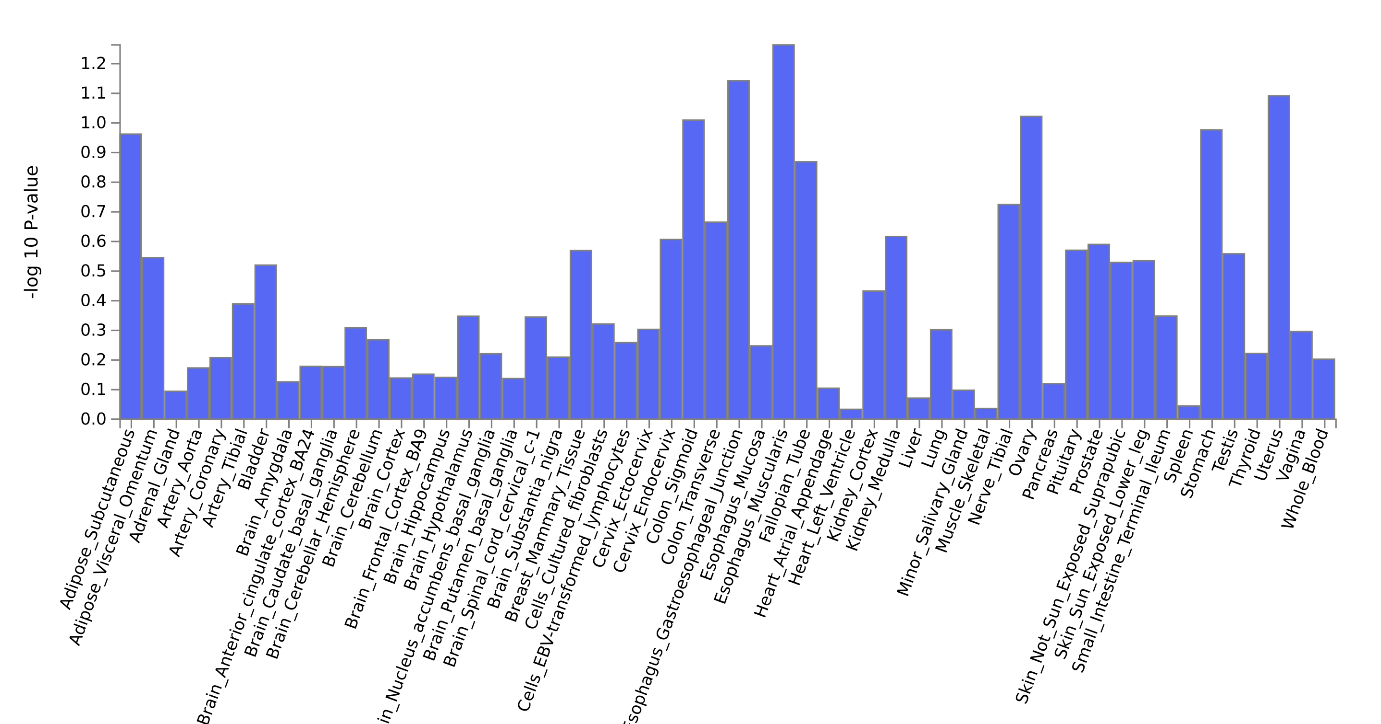

**Supplementary Figure 4.** MAGMA gene-property tissue enrichment analysis (GTEx V8) of GTEx 53 tissues types using the full distribution of SNP p-values from GWIS.

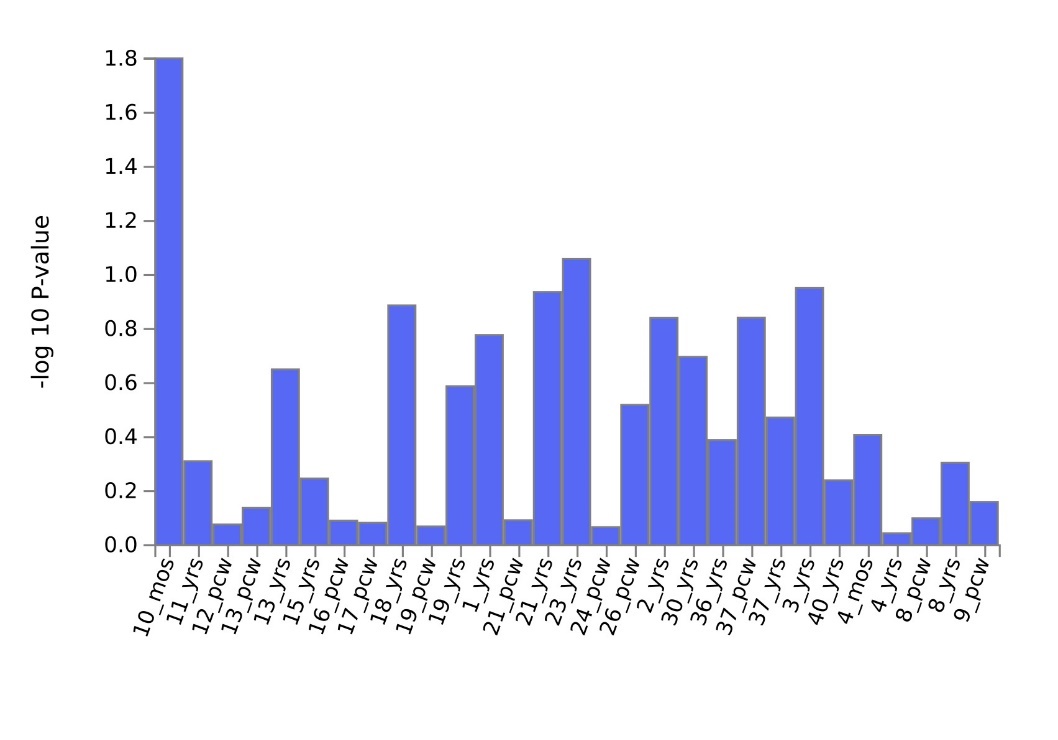

**Supplementary Figure 5.** MAGMA gene-property tissue enrichment analysis (GTEx V8) of BrainSpan 29 ages of brain samples using the full distribution of SNP p-values from GWIS.

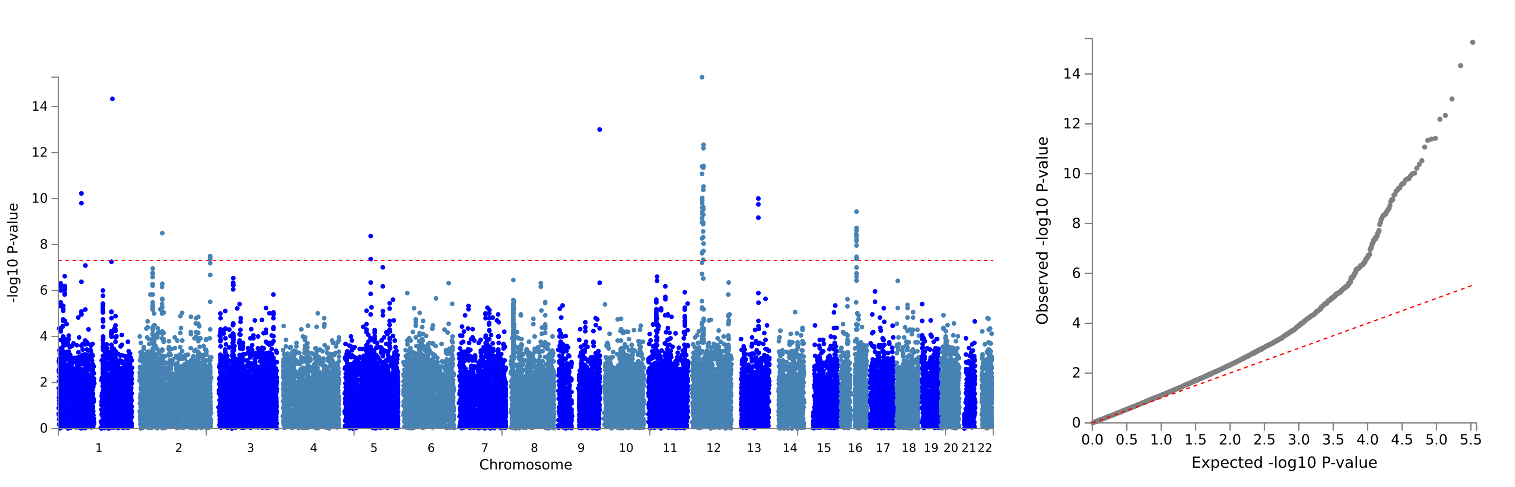

**Supplementary Figure 6.** Manhattan and Q-Q plots for chronotype GWAS among those with low daylight exposure ((≤1.5 h; *n* = 121,922).

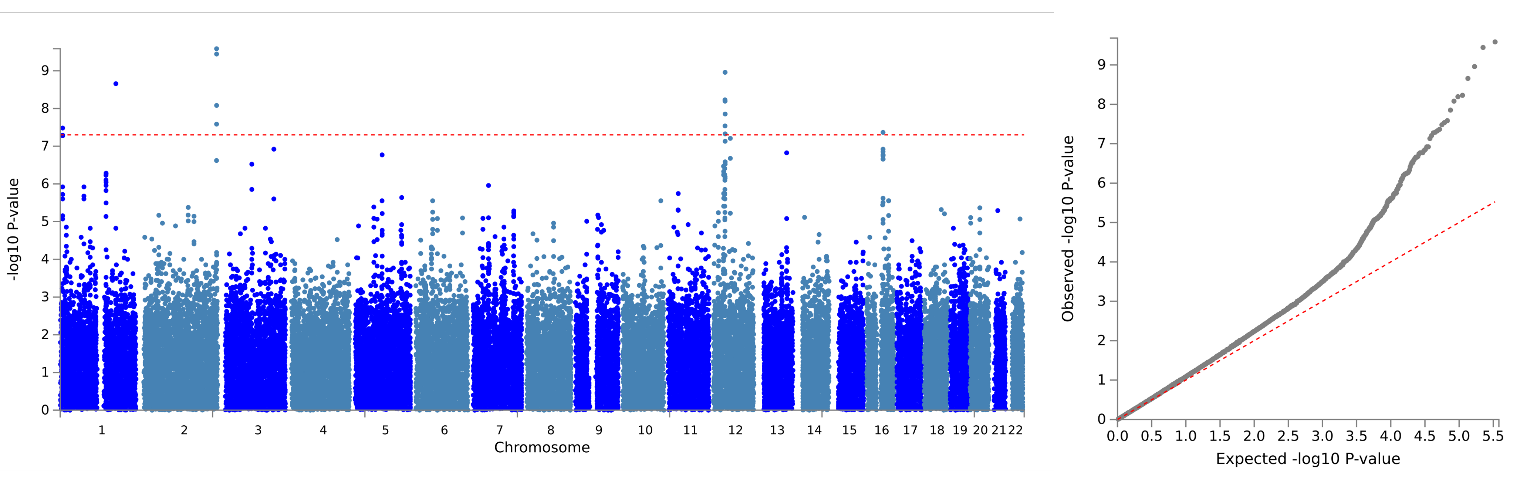

**Supplementary Figure 7.** Manhattan and Q-Q plots for chronotype GWAS among those with low daylight exposure (≥3.5 h; *n* = 118,429).
